# Relationship Between Liver Cancer Mortality and Racial Residential Segregation Across Wisconsin Metropolitan Areas

**DOI:** 10.1101/2020.02.23.20026039

**Authors:** Amin Bemanian, Laura D. Cassidy, Raphael Fraser, Purushottam W. Laud, Kia Saeian, Kirsten M. M. Beyer

**Affiliations:** Institute for Health & Equity, Medical College of Wisconsin, 8701 Watertown Plank Road, Milwaukee, WI 53226; Medical Scientist Training Program, Medical College of Wisconsin, 8701 Watertown Plank Road, Milwaukee, WI 53226; Division of Gastroenterology and Hepatology, Medical College of Wisconsin, 8701 Watertown Plank Road, Milwaukee, WI 53226

## Abstract

**Purpose:** This cross-sectional study tested relationships between racial segregation and liver cancer across several different metropolitan areas in Wisconsin.

**Methods:** Tract level liver cancer mortality rates in Wisconsin were calculated using cases from 2003-2012. Hotspot analysis was conducted and segregation scores in high, low, and baseline mortality tracts were compared with ANOVA. Spatial regression analysis was done controlling for socioeconomic advantage and rurality.

**Results:** Black isolation scores were significantly higher in high mortality tracts compared to baseline and low mortality tracts, but stratification by metropolitan areas found this relationship was driven by two of the five metropolitan areas. Hispanic isolation was predictive for higher mortality in regression analysis, but this effect was not found across all metropolitan areas.

**Conclusions:** This study showed associations between liver cancer mortality and racial segregation, but found this relationship was not generalizable to all metropolitan areas in the study area.

## Introduction

Liver cancer is a major cause of cancer mortality in the United States. In 2016, there were 39,220 newly diagnosed cases of liver cancer and 27,170 liver cancer deaths.^1^ Furthermore, liver cancer has a five-year survival rate of 18% which is exceptionally poor. Liver cancer is notable for having marked racial disparities in incidence and mortality. Black/African Americans (8.4 deaths per 100,000), Hispanics (9.1 per 100,000), Asian/Pacific Islanders (9.5 per 100,000), and American Indians/Alaskan Natives (10.3 per 100,000) have significantly higher mortality rates of liver cancer compared to non-Hispanic Whites (5.7 per 100,000).^2^ Furthermore, Black 5-year liver cancer survival is known to be poorer compared to Whites (13% vs 18%).^1^ Traditionally, these differences have been attributed to racial differences in the prevalence of various risk factors and precursor diseases associated with liver cancer (e.g. viral hepatitis) or due to differences in access to care.^3–5^

Research on other cancers suggests that there is a significant role for the social environment in the development of cancer and its association with racial disparities in cancer outcomes. Ethnic enclaves have been shown to be associated with higher risk of infection-associated cancers such as cervical and liver.^6^ For breast cancer, racial segregation and housing discrimination have been associated with differences in survival.^7–10^ Ethnic enclaves with lower socioeconomic status have also been associated with higher liver cancer incidence rates.^11^ One study of liver cancer prevalence in New York City found that the ZIP code tabulated area (ZCTAs) poverty rate was associated with the prevalence rate of liver cancer even after controlling for the hepatitis C and hepatitis B prevalence rates.^12^ However, compared to other cancers, the literature on the effects of geography and place in liver cancer remains limited. Furthermore, most studies on liver cancer focus on racial disparities for Hispanics and Asians/Pacific Islanders, while Black disparities in liver cancer have been understudied.^6^

Additionally, several studies of racial segregation and cancer have pooled together patients across multiple regions or metropolitan areas to assess if there is a relationship between segregation and cancer risk.^10,13,14^ While this has the benefits of increasing sample size and including a diverse population, there is a risk that the included areas may be too heterogeneous. This is especially significant for studying racial segregation which emerged and manifested in a variety of different ways throughout American history.^15,16^ Furthermore, patterns of immigration in the latter half of the twentieth century have been highly localized by ethnicity to specific areas of the country, and the health impacts of ethnic enclaves have been highly varied.^17–19^ As a result, combining different metropolitan areas may risk obscuring real place-specific relationships.

### Study Purpose

In this paper, we seek to understand how liver cancer in major metropolitan areas in the state of Wisconsin is related to the racial, spatial, and socioeconomic distribution of its residents and how these relationships vary across the state. We focus on two specific questions. First, we seek to identify if there is a relationship between racial residential segregation and liver cancer mortality rates. This analysis is conducted across Wisconsin’s five largest metropolitan areas. Additionally, we seek to determine whether relationships between segregation and liver cancer are place dependent. In order to answer this question, the analysis of segregation and mortality is stratified for each of these five metropolitan areas and multivariate regression models are developed for the two largest Wisconsin metropolitan areas: the Milwaukee-Waukesha-West Allis, WI metropolitan statistical area (MSA) and the Madison, WI MSA.

## Methods

### Mortality Data and Study Area

Tract-level liver cancer standardized mortality rates (SMRs) in Wisconsin were calculated. Mortality data were obtained from the Wisconsin Department of Health Statistics’ Vital Records Service for all cases of liver cancer (ICD10 Codes: C22.0-C22.9) from January 1, 2003 to December 31, 2012 in the state of Wisconsin. A total of 3,204 liver cancer deaths were included in the analysis. Age-adjusted SMRs were calculated using adaptive spatial filtering, which incorporates dynamic spatial filters to overcome issues with having a low number of cases and stabilizes the rates.^21^ This results in a continuous surface which was averaged per tract to estimate tract level rates.

The study area included the five largest metropolitan areas in Wisconsin, excluding any MSAs which crossed state boundaries. These were the Milwaukee-Waukesha-West Allis MSA, the Madison MSA, the Appleton-Oshkosh-Neenah combined statistical area (CSA), the Green Bay MSA, and the Racine MSA. The Appleton-Oshkosh-Neenah combined area was used instead of the Appleton and Oshkosh-Neenah MSAs, because Appleton and Neenah have an interconnected transit system and overlapping municipalities despite being in separate MSAs. Based on the 2010 US census, these five regions combined comprise 3,055,652 residents and 53.7% of Wisconsin residents.^22^ The two largest areas, Milwaukee and Madison, are notable for having very different demographic and socioeconomic characteristics. Maps showing the demographics of these areas are shown in figures 1 and 2. Milwaukee (MSA 2010 pop. 1,555,908) was historically a major manufacturing and industrial hub which had a large influx of African-Americans during the Great Migration in the early 20^th^ century and later experienced a significant exodus of Whites from the city center to the suburbs.^15,23^ These migration patterns resulted in Milwaukee being more racially diverse than the rest of the state (69.0% White, 16.4% Black, 9.5% Hispanic). In a 2010 study of segregation across the 100 largest metropolitan areas in the United States, Milwaukee ranked first in Black-White dissimilarity and ninth in Hispanic-White dissimilarity.^24^ In contrast, Madison (MSA 2010 pop. 568,593) is the state capital and its economy has centered around the state government and the University of Wisconsin, so it has not experienced the same economic pressures as Milwaukee. Madison is less racially diverse than Milwaukee (83.7% White, 4.5% Black, 5.4% Hispanic), and it was ranked 71^st^ for Black-White dissimilarity and 65^th^ for Hispanic-White dissimilarity.^24^

**Figure 1:**
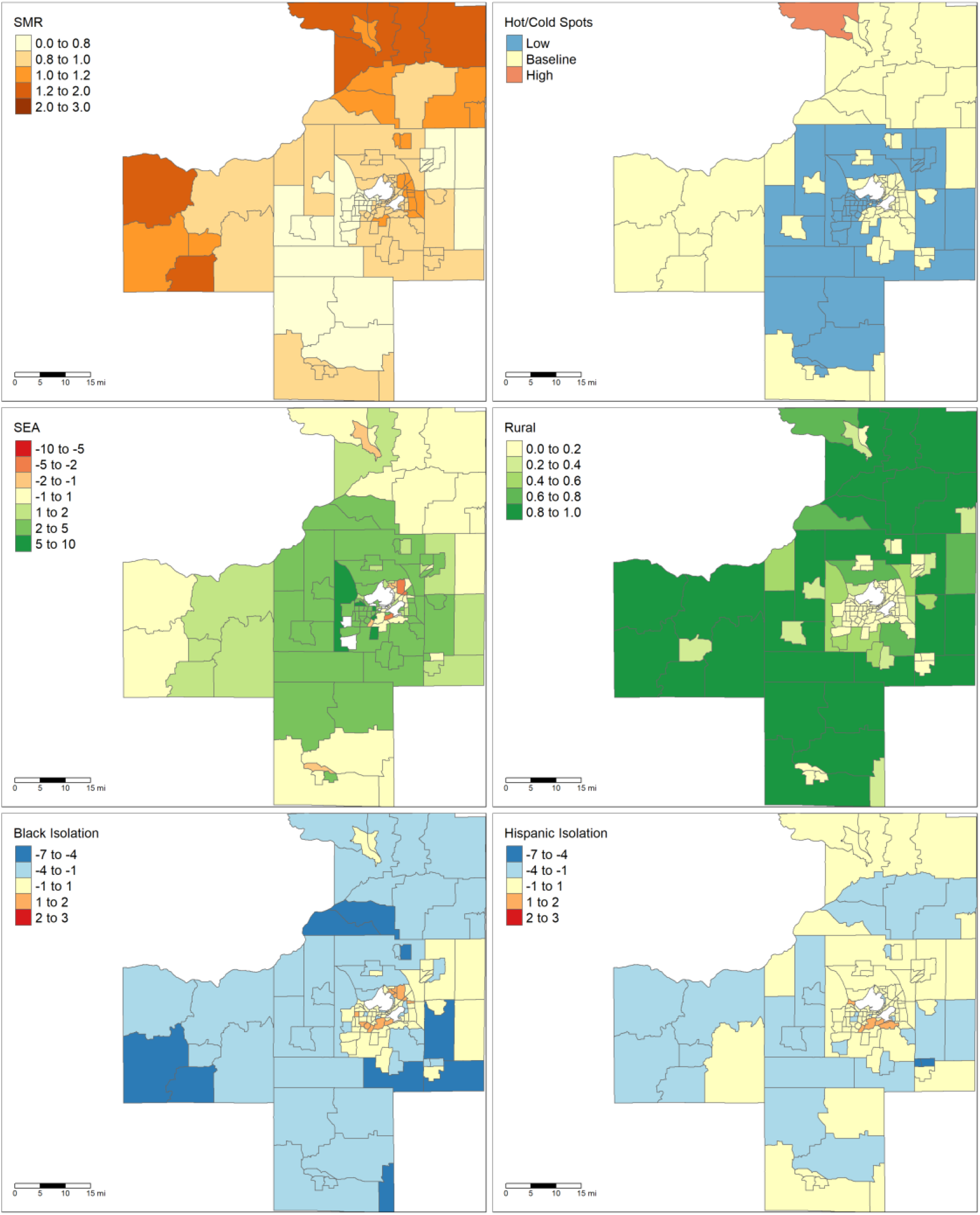
Maps of Madison, WI MSA. Upper right: liver cancer SMRs, upper left: high and low mortality clusters of liver cancer, middle left: socioeconomic advantage index with white tracts indicating areas whose ACS errors were outliers, middle right: rurality, lower left: Black local isolation scores, lower right: Hispanic local isolation scores

**Figure 2:**
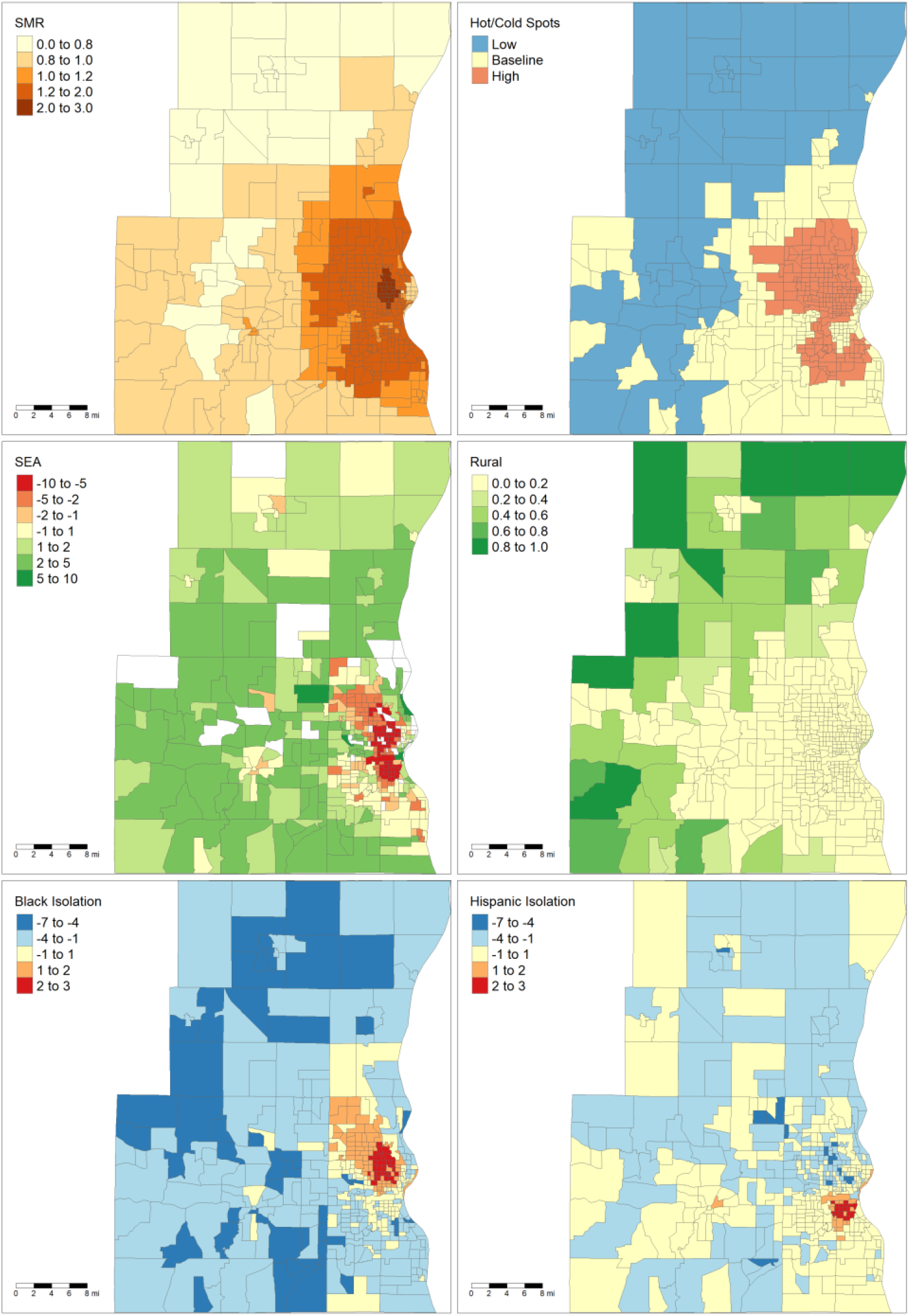
Maps of Milwaukee-West Allis-Wauwatosa, WI MSA. Upper right: liver cancer SMRs, upper left: high and low mortality clusters of liver cancer, middle left: socioeconomic advantage index with white tracts indicating areas whose ACS errors were outliers, middle right: rurality, lower left: Black local isolation scores, lower right: Hispanic local isolation scores

### Demographic Data and Segregation Measurement

Demographic data were obtained from the 2008-2012 five-year American Community Survey. Neighborhood socioeconomic status was calculated using an index of socioeconomic advantage (SEA) which has been previously used for a study of liver cancer mortality in Wisconsin.^20^ This index is calculated from percent of tract residents who graduated with a bachelor’s degree or higher, percent of residents who were unemployed, and the tract’s median household income. Due to concern for instability in the estimates for the socioeconomic variables, SEA was not calculated in tracts if the standard errors for any of the three ACS variables were outliers (> 1.5 the interquartile range). Positive SEA scores indicate higher socioeconomic status relative to the statewide average, and SEA has an approximately normal distribution. Rurality was calculated as the proportion of rural census blocks within a census tract, according to the 2010 US Census urban/rural designations.^25^

Tract-level racial segregation was measured using local Black and Hispanic isolation scores which were calculated using the Local Exposure/Isolation (LEx/Is) index.^9^ This metric measures how much a subarea’s racial/ethnic demographics deviate from the expected case that all racial/ethnic groups are equally distributed across a region. Positive local isolation scores indicate the subarea has a higher proportion of that specific racial/ethnic group than the proportion in the region. LEx/Is scores are scaled using a base-10 logit function.

### Statistical Analysis

Clusters of high and low mortality (i.e. hot and cold spots) were identified using the Getis-Ord Gi statistic.^26^ A threshold of *p* < 0.05 was used to identify high and low clusters. The calculation of SMRs included the entire state in order to have stable enough populations to conduct the ASF technique in the outer tracts of MSAs. As a result, mortality cluster identification involved the entire state as well. Kruskal-Wallis analysis of variance (ANOVA) was used to test differences in segregation, SEA, and rurality across low-mortality, high-mortality, and baseline mortality tracts. ANOVA was conducted across all the study areas and then stratified by each area. Log-linear regression models to predict liver cancer SMRs were developed. These regressions incorporated spatially adjusted errors in order to correct for significant spatial autocorrelation.^27^ The first regression included all five metropolitan areas and used fixed effects to control for baseline differences in SMR across the metropolitan areas. The regression was then repeated for specifically the Milwaukee and Madison MSAs to assess if there were metro-specific relationships. Milwaukee and Madison were selected for individual regression analyses because they were large enough MSAs to not be concerned with overfitting (n of 429 and 131 tracts, respectively). All analysis was conducted using the R statistical programming language.^28^

## Results

Comparisons of the five MSAs are shown in table 1. The average liver cancer SMR and proportion of tracts in a mortality hot or cold spot is significantly different across the MSAs. Milwaukee had the highest mean SMR (1.311) of all MSAs and the largest proportion of tracts in a high mortality cluster (45.2%). The Appleton MSA had significantly lower mortality than the other tracts with a mean SMR of 0.773, no high mortality cluster tracts, and 95.7% of tracts in low mortality clusters. SEA significantly differed across MSAs, with Madison having the highest mean SEA and Milwaukee having the lowest. Milwaukee was significantly more urban than all other MSAs.

**Table 1:** Comparison of metropolitan study areas. APL: Appleton-Oshkosh, GBY: Green Bay, MAD: Madison, MKE: Milwaukee, RAC: Racine. KW H: Kruskal-Wallis one-way analysis of variance H value.

|  | APL |  | GBY |  | MAD |  | MKE |  | RAC |  |  |  |
| --- | --- | --- | --- | --- | --- | --- | --- | --- | --- | --- | --- | --- |
|  | *N* |  | *N* |  | *N* |  | *N* |  | *N* |  |  |  |
| **All Tracts** | 92 |  | 68 |  | 131 |  | 429 |  | 44 |  |  |  |
| **Mortality Clusters** | *n* | % | *n* | % | *n* | % | *n* | % | *n* | % | $\chi^2$ | *P* |
| **High** | 0 | 0.0 | 3 | 4.4 | 1 | 0.7 | 194 | 45.2 | 3 | 6.8 | 393.9 | < **0.001** |
| **Low** | 88 | 95.7 | 12 | 17.6 | 56 | 42.7 | 56 | 13.1 | 7 | 15.9 |  |  |
|  | Mean | SD | Mean | SD | Mean | SD | Mean | SD | Mean | SD | KW *H* | *P* |
| **SMR** | 0.773 | 0.054 | 1.036 | 0.210 | 0.897 | 0.144 | 1.311 | 0.463 | 1.104 | 0.188 | 276.2 | < **0.001** |
| **SEA** | 0.713 | 1.664 | 0.067 | 2.289 | 1.857 | 1.803 | -0.478 | 3.417 | -0.245 | 2.284 | 52.6 | < **0.001** |
| **Black Isolation** | -1.243 | 1.679 | -1.088 | 1.842 | -0.938 | 1.579 | -1.254 | 2.259 | -0.725 | 1.599 | 6.0 | 0.198 |
| **Hispanic Isolation** | -0.391 | 0.984 | -0.731 | 1.447 | -0.466 | 1.032 | -0.744 | 1.352 | -0.406 | 0.911 | 19.9 | < **0.001** |
| **Rural** | 0.214 | 0.349 | 0.288 | 0.396 | 0.279 | 0.377 | 0.065 | 0.183 | 0.171 | 0.260 | 93.7 | < **0.001** |

Boxplots comparing mortality coldspots, mortality hotspots, and baseline mortality tracts are shown in figure 3. These analyses were conducted across all five regions and then stratified for each region. Since Appleton had no mortality hotspots, its analysis was a simple non-parametric comparison of the means with no further multiple comparisons testing. Across all regions, higher Black isolation, lower SEA, and lower rurality were associated with increased likelihood of a tract being in a high mortality cluster and decreased likelihood of being in a low mortality cluster. Hispanic isolation was found to be the highest in the baseline tracts with both high and low mortality cluster tracts having significantly lower Hispanic isolation scores. Stratifying the analysis, we find that only Milwaukee and Racine have evidence of a significant relationship between Black isolation and mortality cluster status. For Milwaukee, there is a significant, monotonic relationship from low to high clusters, but Racine showed no significant difference between average and high clusters. For socioeconomic status, only Milwaukee and Madison showed a significant relationship with cluster status. For rurality, Milwaukee, Racine, and Green Bay showed evidence of the trend found in the pooled analysis. However, in the greater Appleton metropolitan area the low mortality cluster was associated with lower rurality. Notably, all significant relationships in the pooled analysis were also significant in the Milwaukee analysis, and only two significant comparisons in Milwaukee (low vs baseline mortality tracts for Black Isolation and low vs high mortality tracts for Hispanic isolation) were not significant in the pooled analysis.

**Figure 3:**
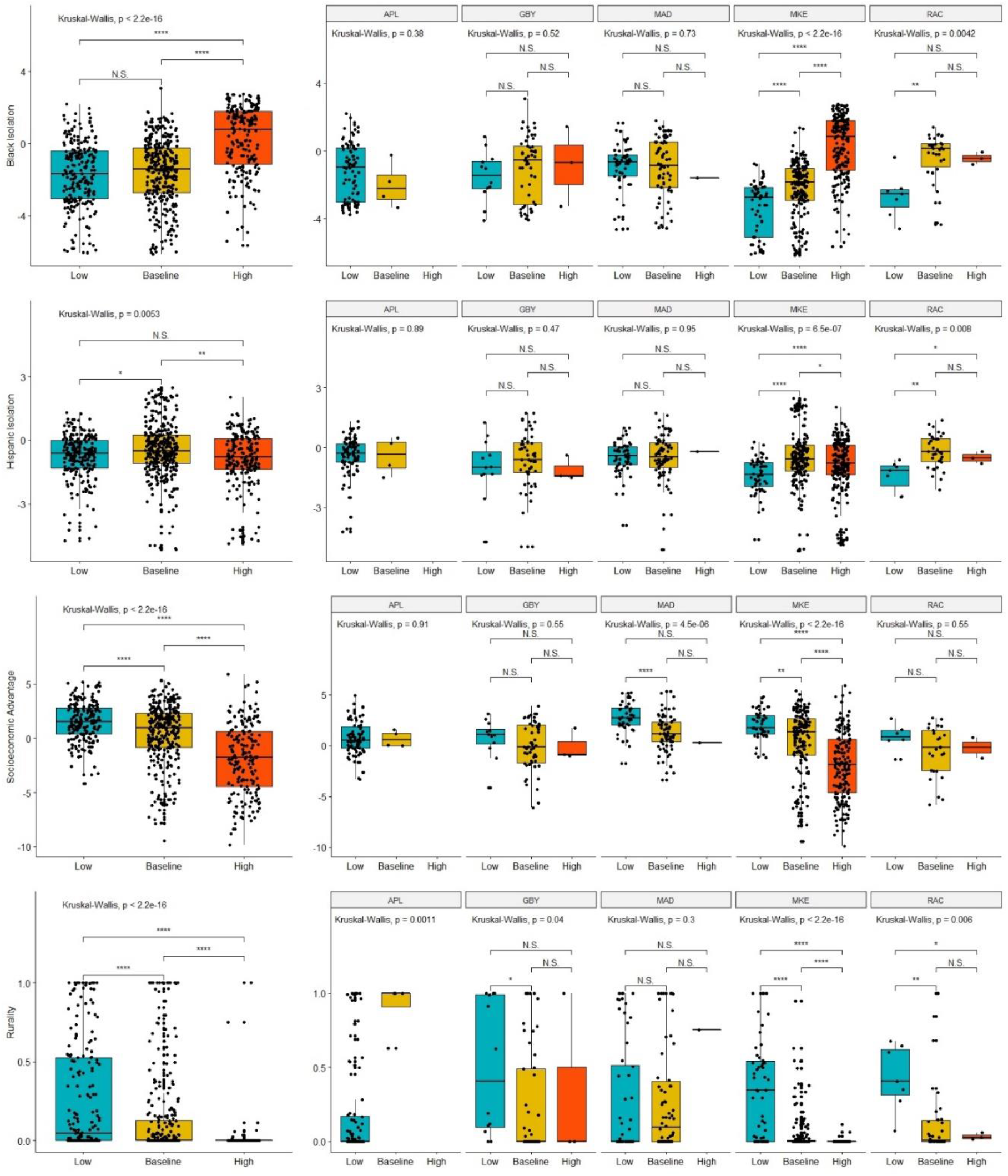
Boxplots comparing segregation and other demographic variables within high, baseline, and low tracts. Left most column indicates comparison across all five MSAs, while other columns indicate comparison within each individual MSA. Differences tested using a Kruskal-Wallis test. Multiple comparison p-values adjusted with Benjamni-Hochberg correction. * indicates p < 0.05, ** indicates p < 0.01, *** indicates p < 0.001, **** indicates p < 0.0001

The log-linear regression models to estimate tract liver cancer SMRs are shown in table 2. This includes one model which pools all five areas and two specific models focusing on the Madison and Milwaukee MSAs. All models were adjusted using spatial errors, and the pooled analysis included hierarchical variables to control for baseline SMR differences for each MSA. Non-spatially adjusted regression was tested as well, but the residuals were significantly spatially autocorrelated (testing with Moran’s I: *p* < 0.001). Racine had a significantly higher baseline SMR with a rate ratio (RR) of 1.581 relative to Appleton. Hispanic isolation was found to be the only other significant predictor of SMR, with higher Hispanic isolation associated with increased SMR. The stratified analysis of Milwaukee also found a similar relationship between Hispanic isolation and liver cancer SMR. No predictors were significant in the Madison model.

**Table 2:** Spatial regression analysis results. First set of columns is the result of the pooled analysis across all five MSAs. Second and third set of columns is result of analysis limited to tracts within the Milwaukee and Madison MSAs respectively. RR: Rate ratio

|  | All MSAs |  | Milwaukee MSA |  | Madison MSA |  |
| --- | --- | --- | --- | --- | --- | --- |
|  | RR | 95% CI | RR | 95% CI | RR | 95% CI |
| **SEA** | 0.997 | 0.992-1.001 | 0.994 | 0.988-1.000 | 0.997 | 0.987-1.008 |
| **Black Isolation** | 1.001 | 0.996-1.006 | 1.004 | 0.996-1.012 | 0.996 | 0.983-1.008 |
| **Hispanic Isolation** | **1.007** | **1.001-1.014** | **1.015** | **1.005-1.024** | 1.006 | 0.990-1.022 |
| **Rural** | 1.005 | 0.967-1.045 | 0.985 | 0.906-1.069 | 0.999 | 0.939-1.064 |
| **MSA Effect** | - | - | - | - | - | - |
| *Appleton* | 1 (Control) | NA | - | - | - | - |
| *Green Bay* | 1.019 | 0.879-1.182 | - | - | - | - |
| *Madison* | 1.182 | 0.699-1.737 | - | - | - | - |
| *Milwaukee* | 1.357 | 0.956-1.927 | - | - | - | - |
| *Racine* | **1.581** | **1.088-2.298** | - | - | - | - |

## Discussion

This paper sought to identify what relationships existed between liver cancer mortality and segregation, and how these relationships vary across different metropolitan areas. Our analysis focused on five different metropolitan areas which, despite being in the same state, are very different in demographics. Milwaukee and Racine tend to be more urban and have lower average socioeconomic status compared to the other regions studied. Furthermore, they had the highest average levels of liver cancer mortality. Given these differences in baseline characteristics, it is not entirely surprising that relationships between demographic variables and cancer mortality varied significantly across MSAs. Comparing the pooled hotspot analyses to the stratified analyses reveals that the pooled findings are primarily being driven by relationships found in the Milwaukee MSA. The population of the Milwaukee MSA is greater than the other four areas combined (1,555,954 vs 1,499,778) and it comprises 56.2% of the number of tracts included in the analysis.^22^ This highlights how one study area can skew the findings of a combined analysis.

The mortality hotspot analyses did show evidence of potential relationships between liver cancer and racial segregation. In the Milwaukee MSA, Black isolation was significantly higher in mortality hotspots and significantly lower in mortality coldspots than in baseline tracts. The only other MSA which had similar findings was Racine. Milwaukee (16.8%) and Racine (11.1%) do have a substantially larger proportion of Black residents compared to the other MSAs (Madison: 4.6%, Green Bay: 1.8%, Appleton-Oshkosh: 1.3%). Interestingly, while Milwaukee has the highest degree of segregation as measured by the Black-White dissimilarity score (0.783), Racine’s level of segregation (0.475) falls in the same range as the other areas (Madison: 0.478, Green Bay: 0.480, Appleton-Oshkosh: 0.417).^29^ Despite the clear relationship between Black isolation and mortality in the hotspot analysis in Milwaukee, the spatially adjusted regression found no relationship. However, the spatially adjusted regression is complicated by the fact that the ASF method used to calculate liver cancer mortality uses cases in neighboring tracts in order to stabilize the rates.^20,21^ Therefore, the liver cancer SMRs are intrinsically spatially dependent. Furthermore, Black isolation is heavily clustered within a single region in Milwaukee (Figure 2). It is hard to determine if there is a consistent relationship between Black isolation and liver cancer mortality or if the relationship is specific to the place itself.

The relationship between Hispanic isolation and liver cancer mortality is more ambiguous. Hotspot analysis found significant differences for Hispanic isolation in Milwaukee, Racine, and the pooled analysis, and the regression analysis showed that Hispanic isolation was associated with increased liver cancer mortality even with spatial adjustment. However, deeper inspection of the hotspot analysis shows that the both mortality hotspots and coldspots have significantly lower Hispanic isolation than the baseline tracts in Milwaukee. The lack of a clear exposure-response relationship for Hispanic isolation makes inferring a meaningful relationship difficult. The pooled hotspot analyses found monotonic exposure-response relationships for socioeconomic advantage and rurality, but neither of these predictors were significant in any of the spatial regression analyses. Given that socioeconomic status and racial demographics of neighborhoods are frequently related, there was concern multicollinearity could be affecting the regression analyses.^14,30^ However, multicollinearity was tested using variance inflation factors, and all combinations were below 2.0, suggesting weak evidence for multicollinearity.

Interestingly, the relationship between rurality and liver cancer mortality was highly specific to the metropolitan area, with more rural tracts more likely to be liver cancer coldspots in Milwaukee, Racine, and Green Bay while less rural tracts were more likely to be coldspots in the Appleton-Oshkosh area. The finding that rurality was associated with decreased mortality in the pooled analysis was surprising given how previous studies have shown that rural counties have higher rates of risk factors associated with liver cancer such as obesity and smoking.^31^ Some of this discrepancy may be due to how rurality was defined in this study. The rurality score represents the proportion of blocks within the census tract that are classified as “rural” by the US Census Bureau. Per the Census Bureau, an urban area is defined as any area of census blocks or tracts which has a core population of at least 2,500 people and any outlying non-residential areas attached to it.^25^ Any census block which falls outside of urban areas is classified as “rural.” Comparing figures 1 and 2 reveals that the distribution of rural blocks is very different in Madison versus Milwaukee. Many of Milwaukee’s highly “rural” tracts are immediately adjacent or one neighbor removed from a predominately urban tract. This suggests that these tracts are simply less developed and most likely have similar access to healthcare resources as their more urban neighbors. While only a small fraction of Milwaukee’s tracts has greater than 80% rurality, the majority of Madison’s tracts by land area have that level of rurality. Furthermore, Madison’s outlying counties only have small islands of urbanicity when compared to the large suburban sprawl of Milwaukee. This highlights how “rural” tracts were heterogenous depending on the study area.

This study is limited by its cross-sectional, ecological design, which prevents causal inference. The outcomes and determinants of health under study were defined at the level of the census tract, while liver cancer occurs on the level of an individual. Furthermore, these models can only examine the relationship between the social determinants and liver cancer at the point of death. Given the often long latency from the initial development of liver disease to liver cancer and death,^32,33^ this is an incomplete assessment of the role social determinants may play in affecting liver cancer disparities. Nevertheless, this study provides important insight in identifying which factors may be influencing these disparities and highlights the importance of taking a place-based approach. The relative rarity of liver of liver cancer is another challenge.

We used adaptive spatial filtering to map liver cancer mortality in Wisconsin, because directly adjusted mortality rates cannot be stably calculated for such a small area as a census tract. This approach can result in a smoothing of the mortality rate, which could lead tracts with extreme rates to be regressed to the means of their neighbors. Nevertheless, this is a necessary adjustment to be able to do an analysis on a sub-county level. An additional limitation is the issue of survey error in the socioeconomic variables used to calculate SEA. While the American Community Survey is a major asset for population health and demographics research, the stability of the estimates for small areas such as tracts has been raised as a potential concern.^34^ By removing the tracts with extreme levels of error, we were able to mitigate some of the risk of unstable estimates while continuing to have a high-resolution analysis. Finally, it is important to note that death certificate data has a risk of imperfect coverage of all liver cancer cases. Previous analysis of death certificates found there is an 87.1% detection rate of liver and intrahepatic bile cancers for ICD-9 coded certificates and a 76.9% detection rate for ICD-10 coded certificates.^35^ While there is a risk that missing liver cancer cases may not be truly randomly distributed across Wisconsin’s population, death certificates provide the highest quality data we have available to conduct these analyses.

This study investigated the relationship between tract-level liver cancer mortality with racial segregation, socioeconomic status, and rurality. Additionally, this study highlighted how these relationships can vary across different metropolitan areas even if they are within the same state. Black local isolation was shown to be a predictor of liver cancer mortality in Milwaukee and Racine, but not in other MSAs. Socioeconomic advantage was associated with lower levels of liver cancer mortality. The variable nature of these relationships across metropolitan areas highlights the importance of place in analyses of cancer disparities. Given how the findings of this study differ from previous ones in describing how ethnic enclaves and racial segregation are associated with liver cancer, further work should be done to characterize the distribution of liver cancer across a diverse set of geographic regions.^5,6,12^ More research needs to be done on the level of the individual to identify how racial segregation affects patients with liver disease and liver cancer. In terms of developing public health interventions to combat liver cancer and reduce disparities, this study suggests that it may be more effective to target specific geographic areas and communities with place-dependent interventions rather than deploying interventions over wide areas.

## Data Availability

Due to concerns for patient privacy, raw cancer mortality data is unable to be shared. Racial segregation and socioeconomic variables are available upon request. All data requests should be directed towards the corresponding author, Amin Bemanian.

## Funding/Acknowledgments

The paper was supported by the Clinical & Translation Science Institute of Southeastern Wisconsin (5TL1TR001437-03) and the National Cancer Institute (F30-CA-216947).

## Ethics Statement

This study was approved by the Medical College of Wisconsin’s Institutional Review Board (IRB). Informed consent was not necessary for this study as it was a retrospective study of already deceased patients. Authors have no financial conflicts of interest to disclose.

## Supplemental Figures

**Appleton**

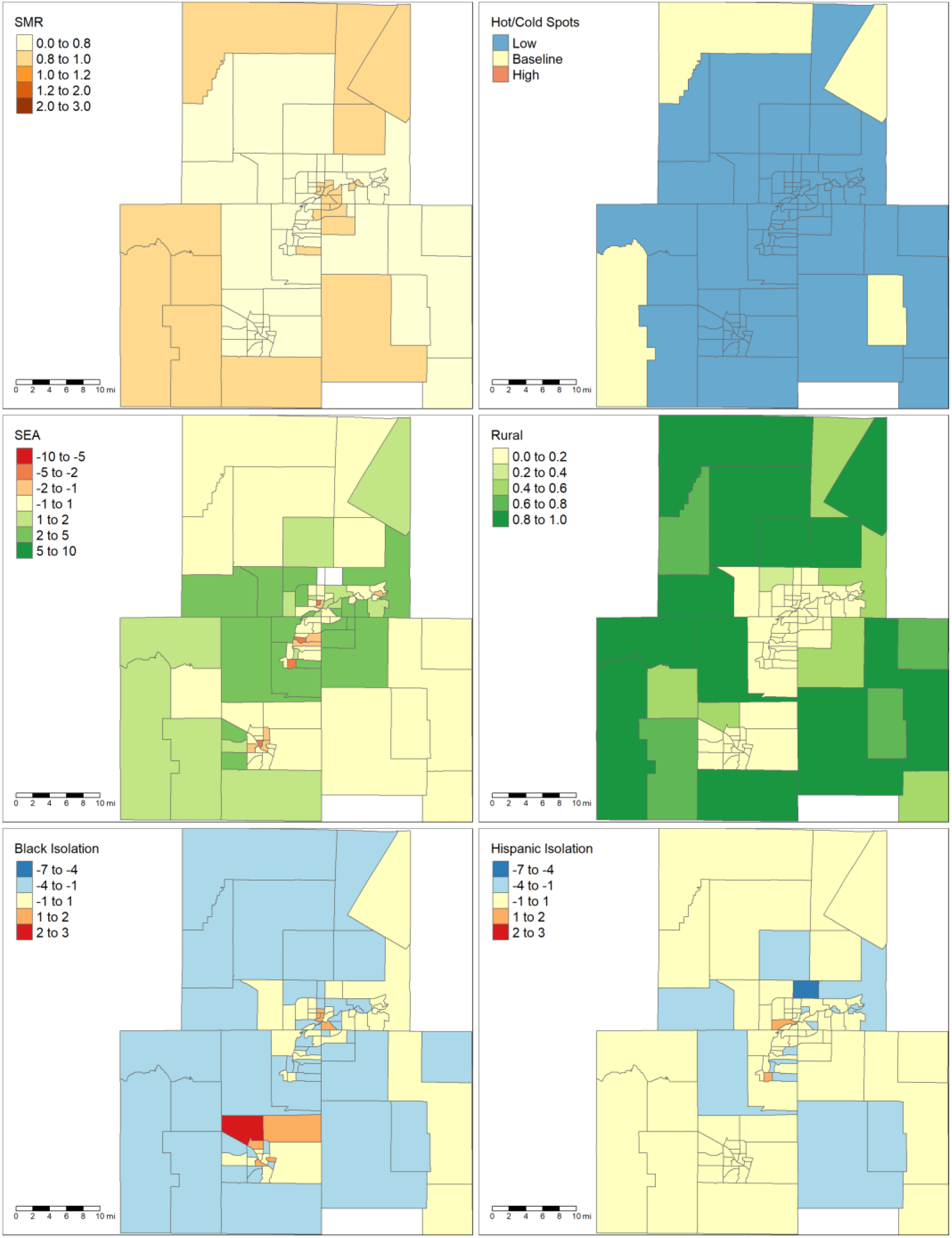

**Green Bay:**

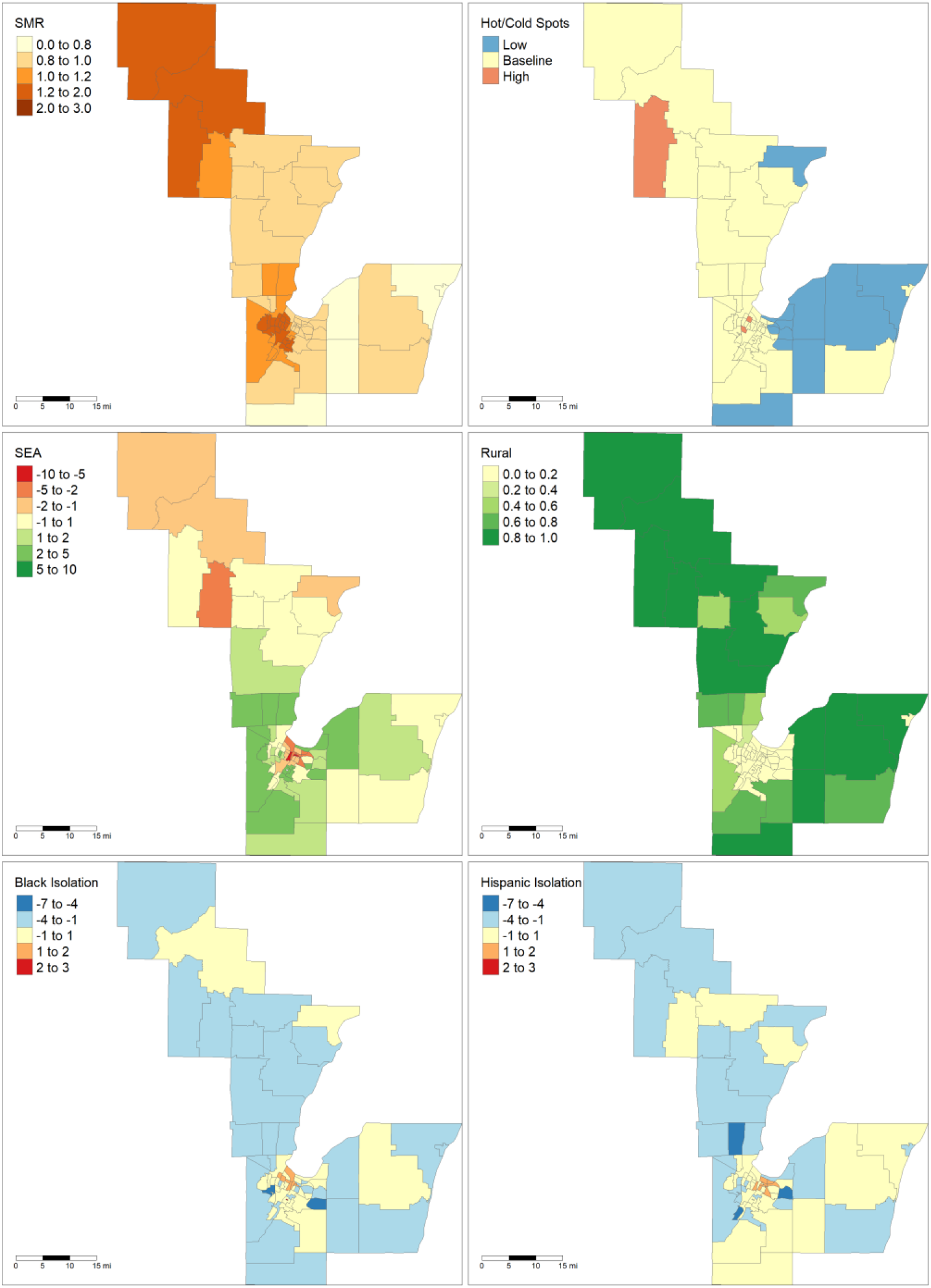

**Racine:**

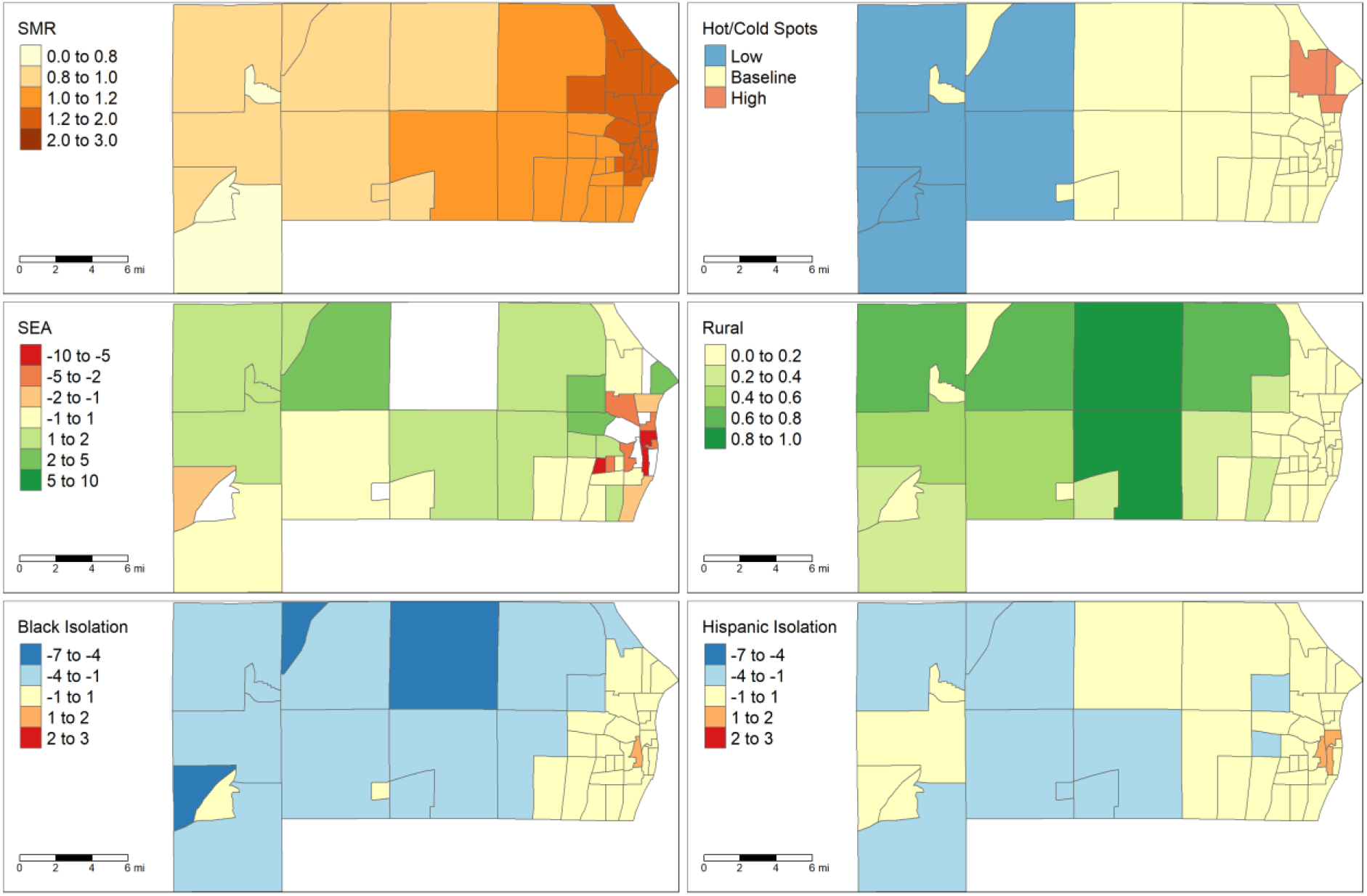

STROBE Statement—Checklist of items that should be included in reports of ***cross-sectional studies***

|  | Item No | Recommendation | Page No |
| --- | --- | --- | --- |
| Title and abstract | 1 | (a) Indicate the study’s design with a commonly used term in the title or the abstract | 1 |
|  |  | (b) Provide in the abstract an informative and balanced summary of what was done and what was found | 1 |
| Introduction |  |  |  |
| Background/rationale | 2 | Explain the scientific background and rationale for the investigation being reported | 2 |
| Objectives | 3 | State specific objectives, including any prespecified hypotheses | 2 |
| Methods |  |  |  |
| Study design | 4 | Present key elements of study design early in the paper | 4 |
| Setting | 5 | Describe the setting, locations, and relevant dates, including periods of recruitment, exposure, follow-up, and data collection | 4 |
| Participants | 6 | (a) Give the eligibility criteria, and the sources and methods of selection of participants | 4 |
| Variables | 7 | Clearly define all outcomes, exposures, predictors, potential confounders, and effect modifiers. Give diagnostic criteria, if applicable | 5 |
| Data sources/<br>measurement | 8* | For each variable of interest, give sources of data and details of methods of assessment (measurement). Describe comparability of assessment methods if there is more than one group | 4 |
| Bias | 9 | Describe any efforts to address potential sources of bias | 11 |
| Study size | 10 | Explain how the study size was arrived at | 5 |
| Quantitative variables | 11 | Explain how quantitative variables were handled in the analyses. If applicable, describe which groupings were chosen and why | 5 |
| Statistical methods | 12 | (a) Describe all statistical methods, including those used to control for confounding | 6 |
|  |  | (b) Describe any methods used to examine subgroups and interactions | 6 |
|  |  | (c) Explain how missing data were addressed | 6 |
|  |  | (d) If applicable, describe analytical methods taking account of sampling strategy | 6 |
|  |  | (e) Describe any sensitivity analyses | NA |
| Results |  |  |  |
| Participants | 13* | (a) Report numbers of individuals at each stage of study—eg numbers potentially eligible, examined for eligibility, confirmed eligible, included in the study, completing follow-up, and analysed | NA |
|  |  | (b) Give reasons for non-participation at each stage | NA |
|  |  | (c) Consider use of a flow diagram | NA |
| Descriptive data | 14* | (a) Give characteristics of study participants (eg demographic, clinical, social) and information on exposures and potential confounders | 7 |
|  |  | (b) Indicate number of participants with missing data for each variable of interest | NA |
| Outcome data | 15* | Report numbers of outcome events or summary measures | 18 |
| Main results | 16 | (a) Give unadjusted estimates and, if applicable, confounder-adjusted estimates and their precision (eg, 95% confidence interval). Make clear which confounders were adjusted for and why they were included | 18 |
|  |  | (b) Report category boundaries when continuous variables were categorized | NA |
|  |  | (c) If relevant, consider translating estimates of relative risk into absolute risk for a meaningful time period | NA |
| Other analyses | 17 | Report other analyses done—eg analyses of subgroups and interactions, and sensitivity analyses | 8 |
| **Discussion** |  |  |  |
| Key results | 18 | Summarise key results with reference to study objectives | 9 |
| Limitations | 19 | Discuss limitations of the study, taking into account sources of potential bias or imprecision. Discuss both direction and magnitude of any potential bias | 11 |
| Interpretation | 20 | Give a cautious overall interpretation of results considering objectives, limitations, multiplicity of analyses, results from similar studies, and other relevant evidence | 10 |
| Generalisability | 21 | Discuss the generalisability (external validity) of the study results | 12 |
| **Other information** |  |  |  |
| Funding | 22 | Give the source of funding and the role of the funders for the present study and, if applicable, for the original study on which the present article is based | 1 |

\*Give information separately for exposed and unexposed groups.
**Note:** An Explanation and Elaboration article discusses each checklist item and gives methodological background and published examples of transparent reporting. The STROBE checklist is best used in conjunction with this article (freely available on the Web sites of PLoS Medicine at http://www.plosmedicine.org/, Annals of Internal Medicine at http://www.annals.org/, and Epidemiology at http://www.epidem.com/). Information on the STROBE Initiative is available at www.strobe-statement.org.

